# Development of a Deep Learning Based Framework for Classification of Indian Venomous Snakes Integrated with Explainable Artificial Intelligence for primary and emergency care providers

**DOI:** 10.64898/2026.03.16.26348471

**Authors:** Ikhlaas Ifthikar Abusayeed Manna, Usha Wagle, Badhrinarayanan Balaji, Vrinda Lath, Niranjana Sampathila, Upadya P Sudhakara, Freston Marc Sirur

## Abstract

**Background:** Snakebite envenoming is a significant global health crisis that has been long neglected as a global health priority. It is a huge problem for rural communities of low and middle-income countries, India accounts for the largest proportion of snakebite deaths globally. Timely identification of venomous snakebite and its syndromic pattern is essential for effective administration of antivenom and supportive treatment. Expert identification of snake species and syndromes is not always available in peripheral healthcare settings. This leads to delays, unnecessary referrals, or improper treatment choices. Additionally, diverse snake species distribution and venom variations across regions pose challenges. AI-powered image classification methods can help overcome these barriers. We propose a clinically oriented deep learning pipeline for binary classification of venomous and non-venomous snake species of India using real-world imagery data. This pipeline would serve as a baseline step towards aiding snakebite management at peripheral healthcare setups with scarce resources.

**Methods:** The selected dataset consisted of 20 medically important Indian species. MobileViT-S, ConvNeXt-Tiny, EfficientNet-V2-S and ResNeXt-50 (32×4d) were trained under same conditions for comparison of results. Model interpretability was evaluated using Grad-CAM ++ to ensure that classification was not performed based on background but on features like head shape and stripes present on body. For reliable implementation we connected it to a web interface with human in loop expert verification. Experts can confirm or override predictions in real time.

**Results:** Among the evaluated architectures, ResNeXt-50 (32×4d) showed the most reliable and consistent performance in classifying venomous and non-venomous snakes. It achieved the highest test accuracy, sensitivity, specificity, and F1-score. The model also had strong discriminative ability, with a ROC-AUC of 0.9950 and PR-AUC of 0.9959. These results indicate dependable performance in safety-critical screening situations. Grad-CAM++ visualizations confirmed that predictions were based on anatomically relevant features, especially in the head and body contour areas. This supports model interpretability and reduces background bias.

**Conclusions:** Although the dataset size and single-institution source limit how widely the results can be applied, the proposed framework shows that it’s possible to create a clinically oriented, ready-to-use deep learning system for snakebite triage support. This system is intended as a scalable tool to help rural healthcare workers, emergency responders, and telemedicine platforms in areas where snakebites are common.

**Author Summary:** Snakebite is a major public health concern that disproportionally affects the rural population. Delays in identifying whether a snake is venomous often lead to delayed treatment, unnecessary use of antivenom, or inappropriate referrals. In many rural settings, access to expert snake identification is limited. To address this gap, authors have developed an artificial intelligence (AI)–based image classification system that distinguishes snakes into two clinically relevant categories: venomous or non-venomous. Unlike many previous studies that focused on ideal, high-quality wildlife images, our model was trained using real-world photographs captured in emergency situations, including images taken by patients and field responders under variable lighting and background conditions. This approach improves the model’s relevance to practical healthcare settings. The system achieved high accuracy and was further strengthened by visual interpretability tools and expert verification to ensure reliability. By combining AI-assisted classification with human oversight, this work provides a scalable decision-support tool that may improve early triage, rational antivenom use, and surveillance in snakebite-endemic regions

## 1. Introduction

Snakebite envenoming is a serious and often ignored global health issue [1,2] affecting around 1.8 to 2.7 million people each year. It causes between 81,000 and 138,000 deaths, along with about 400,000 cases of permanent disability worldwide [1,3] In response to this burden, the Government of India launched the National Action Plan for Prevention and Control of Snakebite Envenoming (NASPE). This plan details a strategy that includes capacity building, community awareness, and improved access to antivenom. The goal is to cut snakebite-related deaths and illnesses by 50% by 2030. [4]. The burden falls heavily on disadvantaged rural populations. They have limited access to formal healthcare services. There are issues with first aid response, uneven supplies of antivenom, and insufficient training for medical staff all of which contribute to significant underreporting.[1] Beyond mortality, snakebite has serious socio-economic effects. It causes disability, loss of income, and high health care costs. These issues deepen poverty. [4,5]. India has the largest share of this burden. It has the highest number of snake bite cases and snake bite related deaths.[7] Areas with the most incidents often overlap with regions that have intensive agricultural and plantation activities, along with limited healthcare facilities [8] These factors make India a key area for public health efforts. It is also an urgent place to test technologies that can reduce delays and improve clinical decision making after a snakebite incident.

Effective clinical management of snakebites requires proper species identification and antivenom administration. In India, clinical practice has long focused on the "Big Four": the Spectacled Cobra (*Naja naja*), Common Krait (*Bungarus caeruleus*), Russell’s viper *(Daboia russelii*), and saw-scaled viper (*Echis carinatus*). However, new herpetological and clinical evidence shows that this framework is an oversimplification. Several other important species can cause envenoming, and misidentifying morphologically similar species can lead to incorrect treatment.[9,10] In Coastal Karnataka, apart from the Big 4, there are other medically important species. These include the Hump-nosed pit viper, Malabar pit viper, and King cobra, for which Indian polyvalent ASV provides little to no benefit. Hump-nosed pit vipers (*Hypnale hypnale*) are often mistaken for saw-scaled vipers in Kerala due to their morphological resemblance. This confusion can lead to wrong treatment choices. Additionally, other significant species like the monocled cobra (*Naja kaouthia*) in northeastern India, various pit viper types, and regional variants such as Sochurek’s saw-scaled viper (*Echis carinatus sochureki*) are inadequately addressed by the existing polyvalent antivenoms. Therefore, improving species identification is not just a theoretical issue; it has real effects on treatment. Artificial intelligence can serve as a valuable decision-support tool in snakebite management, particularly where expert identification is unavailable and rapid decisions are required where the expert identification is lacking and quick decisions are needed.

Artificial intelligence (AI) offers a helpful decision-support method when experts are hard to find and quick triage decisions are necessary. Deep learning image models can learn unique patterns from sets of photos, allowing for automatic identification even in changing field conditions.[11]In rural India, where snakebite cases are common and specialist help is scarce, AI-assisted classification systems could help with early risk assessment.[11] The current literature mainly focuses on identifying species using well-curated, high-quality image datasets that are often taken in controlled environments. These images do not accurately represent real-life snakebite situations, where the images tend to be partial, blurred, poorly lit, or taken during emergencies. Furthermore, existing studies typically view snake identification as a wildlife or biodiversity issue, rather than a problem that supports clinical decisions. Research on Indian snake species is also limited, even though India faces the highest global mortality rates from snakebites. The binary distinction between venomous and non-venomous snakes is not adequately covered in the literature, even though this information is crucial for clinical triage. Issues related to class imbalance, practical deployment in resource-limited settings, and integration into real-time clinical workflows also remain underexplored.

Relatively few studies have specifically addressed binary venom classification of Indian snake species within a clinically oriented framework. In this study, we propose an optimized deep learning–based classification framework with the following key contributions:

- Indian snake image species organized into binary classes venomous and Non-venomous for training pre-trained Models.
- Integration of Grad-CAM based explainability techniques improves model interpretability. This ensures that classification decisions are based on anatomically meaningful morphological features.
- Presentation of a clear comparison of venomous classification performance based on performance parameters of the models.

## 2. Methods

### 2.1 Study setting and Image acquisition

The dataset was developed at the Center for Wilderness Medicine, Kasturba Medical College & Hospital, MAHE, Manipal. Multi-source image acquisition was employed to enhance model robustness and ecological validity. Images encountered in real-world clinical settings are frequently captured under suboptimal conditions, including poor lighting, motion blur, partial visibility, complex backgrounds, and variable camera quality. Training the model on images derived from emergency department submissions, roadkill encounters, and field expeditions allowed exposure to diverse environmental contexts and imaging conditions. Such variability reduces the risk of overfitting controlled or idealized image features and improves generalization performance across heterogeneous deployment scenarios.

The primary source of images consisted of photographic evidence provided by patients or patient attenders presenting to the emergency department with a history of snakebite. The images of the culprit species were recorded as part of evidence captured for the VENOMS registry. The VENOMS Registry is a prospective, IEC approved and Clinical Trial Registry-India registered Registry initiated in the year 2019, which systematically records cases of envenomation including snakes, Hymenoptera and marine envenomation. These images were typically captured at the site of the incident by the patients or their family using mobile devices and reflect real-world field conditions.

When live or deceased specimens were brought to the emergency department, the authors obtained high-resolution photographs under controlled conditions to ensure accurate documentation of morphological features. The dataset included images of road-kill specimens encountered during the field activities. Roadkill images provided additional opportunities for full body documentation under natural environment conditions and contributed to species diversity within the dataset.

Additionally, supplementary images were sourced from various field expeditions, herpetological surveys and conflict specimens across India in which the authors (FMS, VL) participated. These expeditions provided authenticated photographic documentation of the wild specimens across varied ecological setting in India.

### 2.2. Species selection & Characteristics

#### The dataset included images of 20 Indian snake species

Bamboo Pit Viper, Beddome’s Keelback, Buff-striped Keelback, Cat Snake, Checkered Keelback, Common Krait, Common Wolf Snake, Green Vine Snake, Hump-nosed Pit Viper, Indian Rock Python, King Cobra, Malabar Pit Viper, Ornate Flying Snake, Rat Snake, Russell’s Kukri, Russell’s Viper, Saw-scaled Viper, Sea Krait, Spectacled Cobra, and Whitaker’s Boa, as shown in Fig 1. These species were selected based on clinical relevance, epidemiology, and frequency of misidentification in emergency settings. The entire dataset comprised images of both intact and partially mutilated snake specimens. In total, it included 1,110 images of venomous snakes and 384 images of non-venomous snakes.

**Fig 1:**
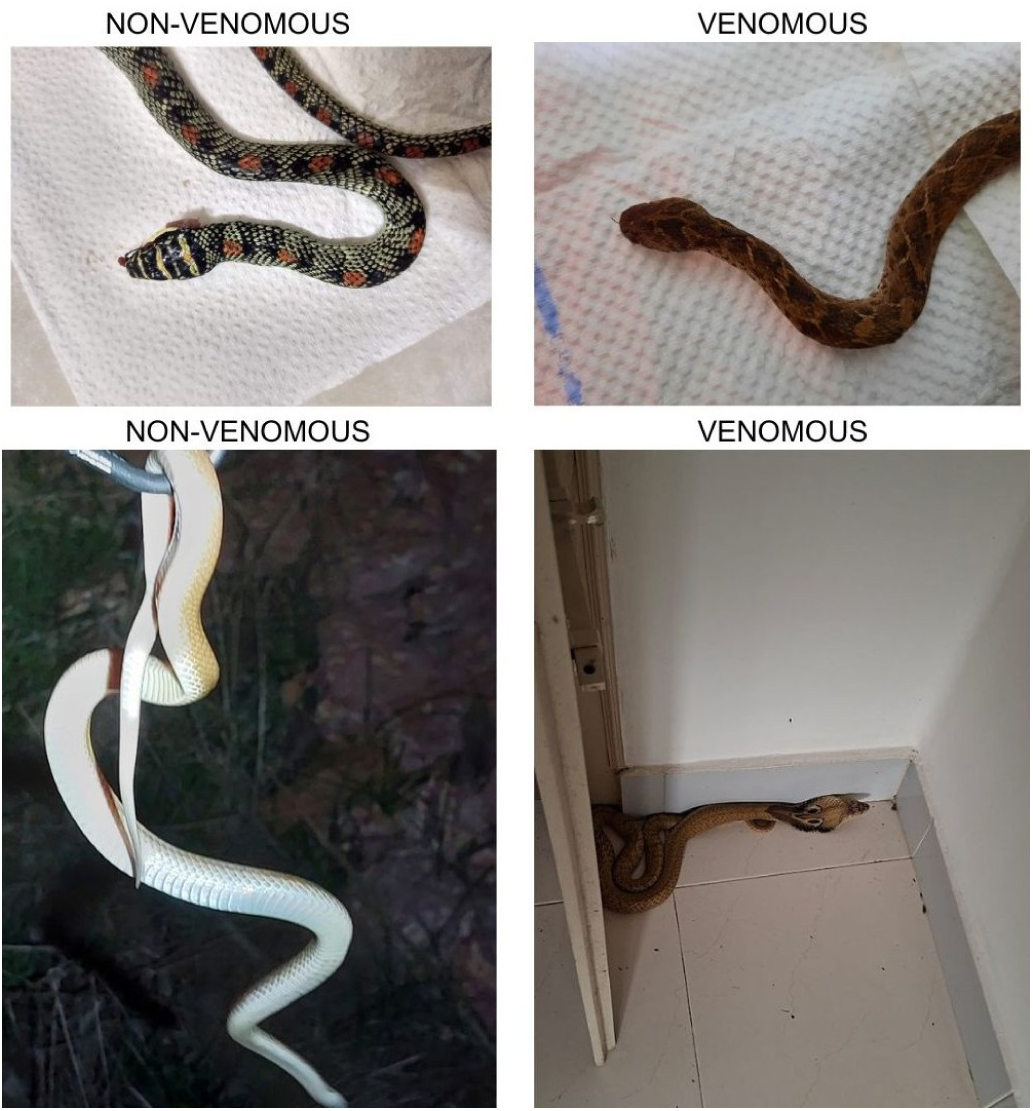
Venomous and Non-Venomous Snake Images Captured with Different sizes in India

### 2.3 Data annotation and categorization

All images were reviewed and annotated by experts with experience in clinical snakebite management and field expeditions. Dataset was subsequently categorized into binary classes Venomous and Non-Venomous, to support model training.

### 2.4 Workflow Diagram

As shown in Fig 2, the end-to-end pipeline for classification of snakes into venomous and non-venomous categories. Raw field images captured under diverse real-world conditions are first subjected to data augmentation, including rotation, flipping, affine transformations, perspective distortion, random resizing, and color jittering, to enhance robustness and reduce overfitting. Augmented images are then processed by selected deep learning architectures (MobileViT-S,[12] ConvNeXt-Tiny[13], EfficientNetV2-S[14], ResNeXt-50[15]), which are trained on the dataset to perform classification. Model interpretability is provided using Grad-CAM++, highlighting image regions contributing most to predictions. [16]The final output categorizes each image as venomous or non-venomous, supporting rapid decision-making in snakebite-prone regions.

**Fig 2:**
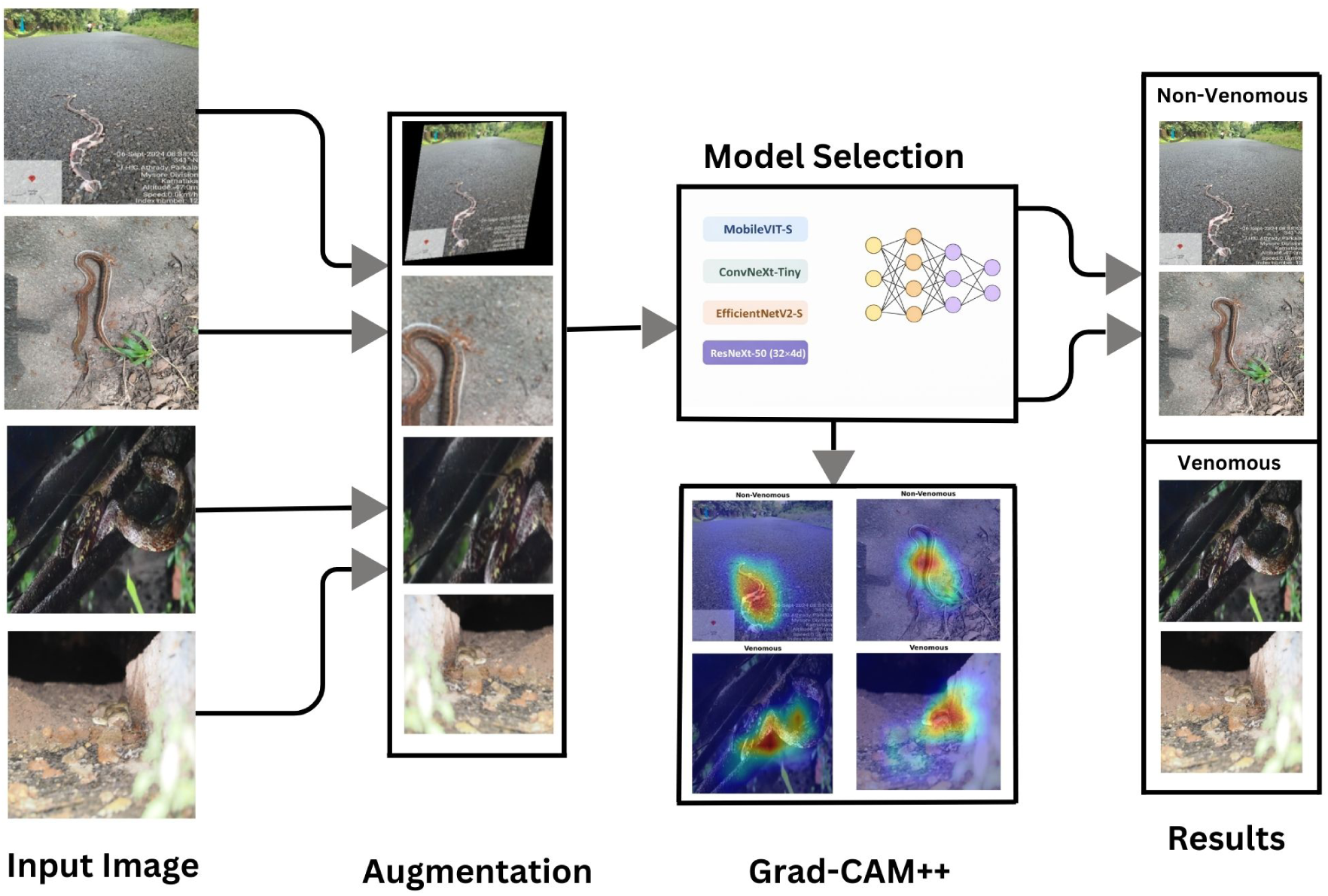
Deep learning framework for detecting Venomous class from Indian Snake images

### 2.5. Data augmentation

Data augmentation (Fig 3) was applied to improve the model’s generalization, as real-world snake images vary widely in orientation, lighting, background, and perspective. To enhance invariance to these variations, geometric and photometric transformations were used. [17] All images were resized to 320 × 320 pixels. The training pipeline included random rotations (±45°), horizontal (50%) and vertical (30%) flipping, affine transformations (shear ±10°), perspective distortion (scale 0.2), and random resized cropping (scale 0.6–1.2) to simulate viewpoint and scale changes [17]. Color jittering modified brightness, contrast, and saturation (±40%) and hue (±15%) to handle lighting variability [17] For validation and testing, images were only resized to 320 × 320 pixels and normalized using the ImageNet mean and standard deviation [18]

**Fig 3:**
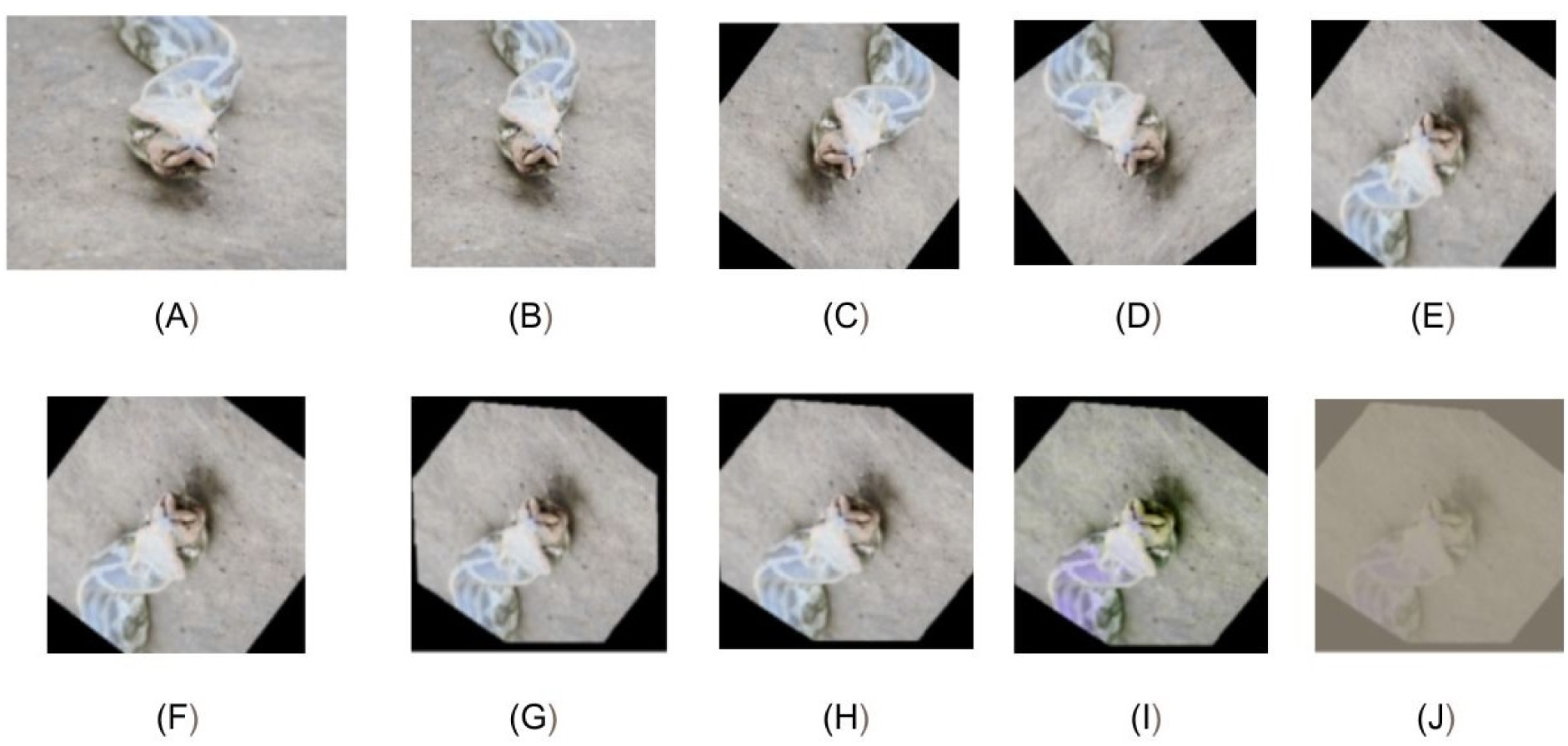
Augmentation Operations on snake data (a) Original Image (b)Resize (c) Random Rotation (d) Horizontal Flip (e) Vertical Flip (f) Affine shear (g) Perspective (h)Random Resize (i)Color Jitter (j) tensor

### 2.6. Model Selection

To identify the most suitable architecture, four models MobileViT-S, [12] EfficientNetV2-S[14] ConvNeXt-Tiny [13] and ResNeXt-50 (32×4d) [15] were trained under identical hyperparameters, data augmentation strategies, and optimization settings to ensure a fair and controlled comparison.

## 3 Results

The training and validation curves indicate stable convergence and good generalization for all architecture as shown in Fig 4. MobileViT-S shows rapid early accuracy gains with closely matched training and validation curves. The loss stabilizes quickly. ConvNeXt-Tiny improves steadily, although it experiences slight loss fluctuations in the later epochs. EfficientNetV2-S has the smoothest and most gradual convergence, with loss consistently decreasing. ResNeXt-50 converges quickly and shows competitive accuracy, experiencing mild validation loss fluctuations before stabilizing. Final model selection was based on the quantitative evaluation metrics described in Fig 5.

**Fig 4:**
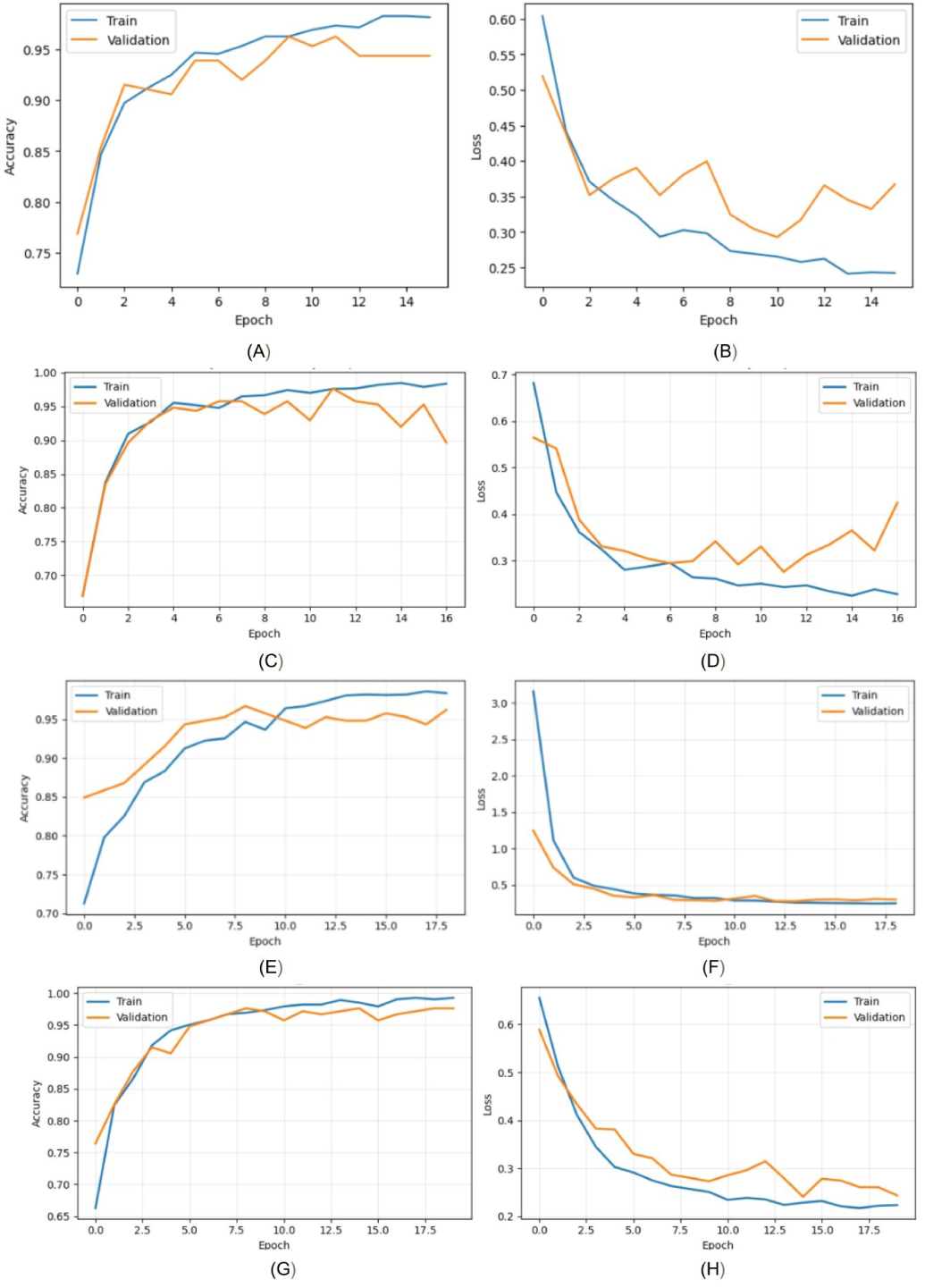
Accuracy and loss curves for trained models - (A–B) MobileViT-S, (C–D) ConvNeXt-Tiny, (E–F) EfficientNetV2-S, and (G–H) ResNeXt-50 (32×4d). Graphs (A, C, E, G) show training and validation accuracy; Graphs (B, D, F, H) show corresponding loss curves.

**Fig 5:**
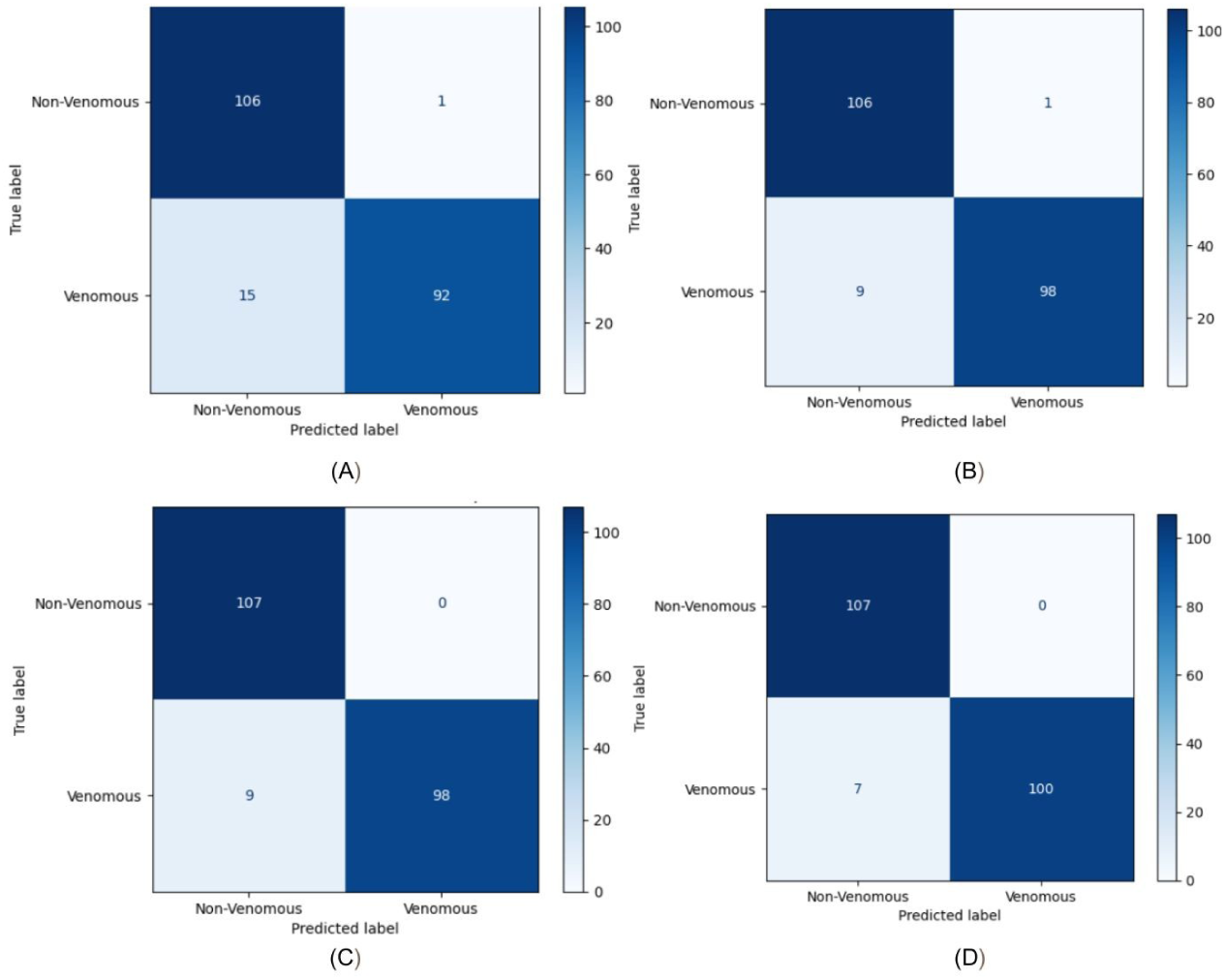
Confusion Metrics - (A) MobileViT-S Metrics,5(B) ConvNeXt-Tiny Metrics,5(C) EfficientNet Metrics,5(D) ResNeXt-50 Metrics

The confusion matrices (Fig. 5) show clear differences in performance across classes, especially for the important venomous class. MobileViT-S correctly classifies 106 non-venomous samples, but it misclassifies 15 venomous cases, which shows lower sensitivity. ConvNeXt-Tiny and EfficientNetV2-S lower the venomous false negatives to 9 while keeping high accuracy for non-venomous samples (106 correct, 1 incorrect), indicating better class balance. ResNeXt-50 performs the best, correctly identifying all 107 non-venomous samples with no false positives and reducing venomous false negatives to 7, which is the lowest among all models. This steady decrease in venomous misclassification highlights how architectural design impacts safety-critical detection. In conclusion, ResNeXt-50 provides the best performance for both class-wise and overall results, making it the best model for the binary snake classification task.

Overall, although all models achieve high predictive accuracy, ResNeXt-50 consistently attains the strongest class-wise performance and the highest overall evaluation metrics under the present experimental conditions, making it the most suitable architecture for the proposed binary snake classification task.

As shown in Table 1, there is a comparison of the four models based on key performance metrics, revealing clear differences in generalization and diagnostic reliability. While MobileViT-S reaches high training accuracy and specificity, its lower sensitivity means it may miss venomous cases, making it less suitable for clinical screening. ConvNeXt-Tiny improves sensitivity with a strong F1 score; however, its relatively low validation accuracy raises concerns about its stability in generalization. EfficientNet-V2-S shows more balanced performance, matching the same sensitivity while achieving perfect specificity and higher validation accuracy, which indicates better convergence behavior. Overall, ResNeXt-50 provides the strongest and most consistent results across all metrics. It has the highest test accuracy, sensitivity, specificity, and F1 score, showing it offers the most reliable performance among the models evaluated.

**Table 1:** Comparative Metrics for Various Models.

| Model name | Train<br>accuracy | Validation<br>accuracy | Test<br>accuracy | Sensitivity | Specificity | F1 score |
| --- | --- | --- | --- | --- | --- | --- |
| MobileVit | 98.12 | 94.32 | 92.52 | 85.98% | 99.07% | 92.49 |
| ConvNext-Tiny | 98.35 | 89.62 | 95.33 | 91.59% | 99.07% | 95.32 |
| EfficientNet-v2-s | 98.35 | 96.23 | 95.79 | 91.59% | 100% | 95.79 |
| ResNeXt-50 | 99.29 | 97.64 | 96.73 | 93.46% | 100% | 96.73 |

The performance of ResNeXt-50 is further validated by the ROC and Precision-Recall curves in Fig 6. The ROC curve reaches an AUC of 0.9950. This shows a strong ability to differentiate between venomous and non-venomous classes. The curve closely follows the top-left corner and significantly surpasses the random classifier baseline. The Precision-Recall curve also shows a PR-AUC of 0.9959. This confirms that the model keeps high precision even when recall levels are high. This is important in safety-sensitive screening since it shows that the model can accurately identify most venomous cases while avoiding many false alarms. Together, these curves support the sensitivity, specificity, and F1 metrics in Table 1. They establish ResNeXt-50 as a strong and dependable classifier for safety-critical snakebite triage applications.

**Fig 6:**
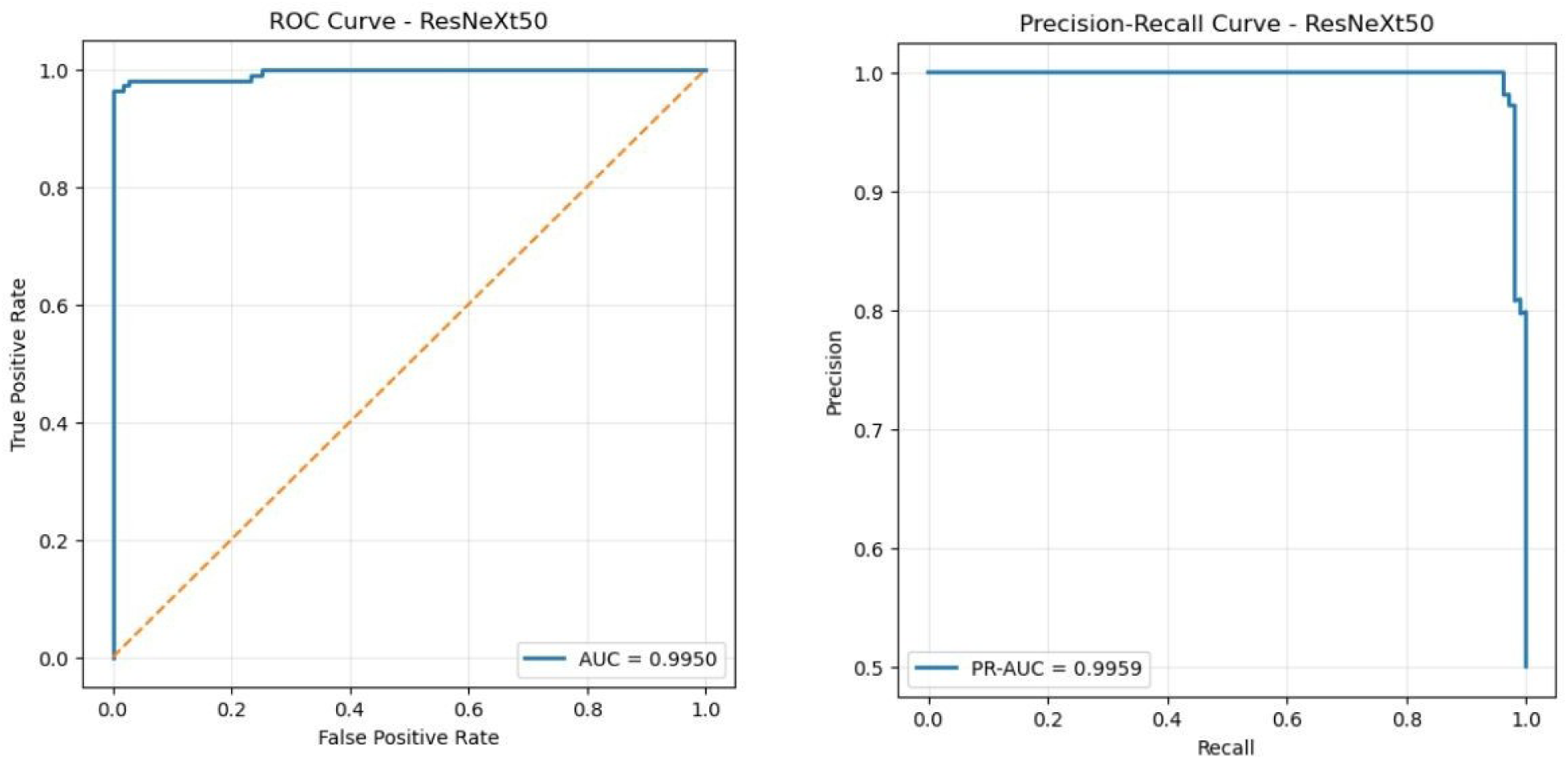
Precision-Recall Curve and ROC Curve for ResNeXt-50 (32×4d) on the test set.

As illustrated in Fig 7(A–C) a representative non-venomous case is analyzed using Grad-CAM++ visualization.[16] As Fig 7(A) shows the original input image, while Fig. 7(B) includes the activation map, highlighting strong responses along the snake’s long body shape indicating that the model attends to anatomically relevant regions .As shown in Fig 7(C) overlays the heatmap onto the original image. This confirms that the model focuses on physical features like body segments and outline patterns. This specific and clear activation suggests that the model uses meaningful, detailed anatomical features for classifying non-venomous snakes while reducing contextual bias

**Fig 7:**
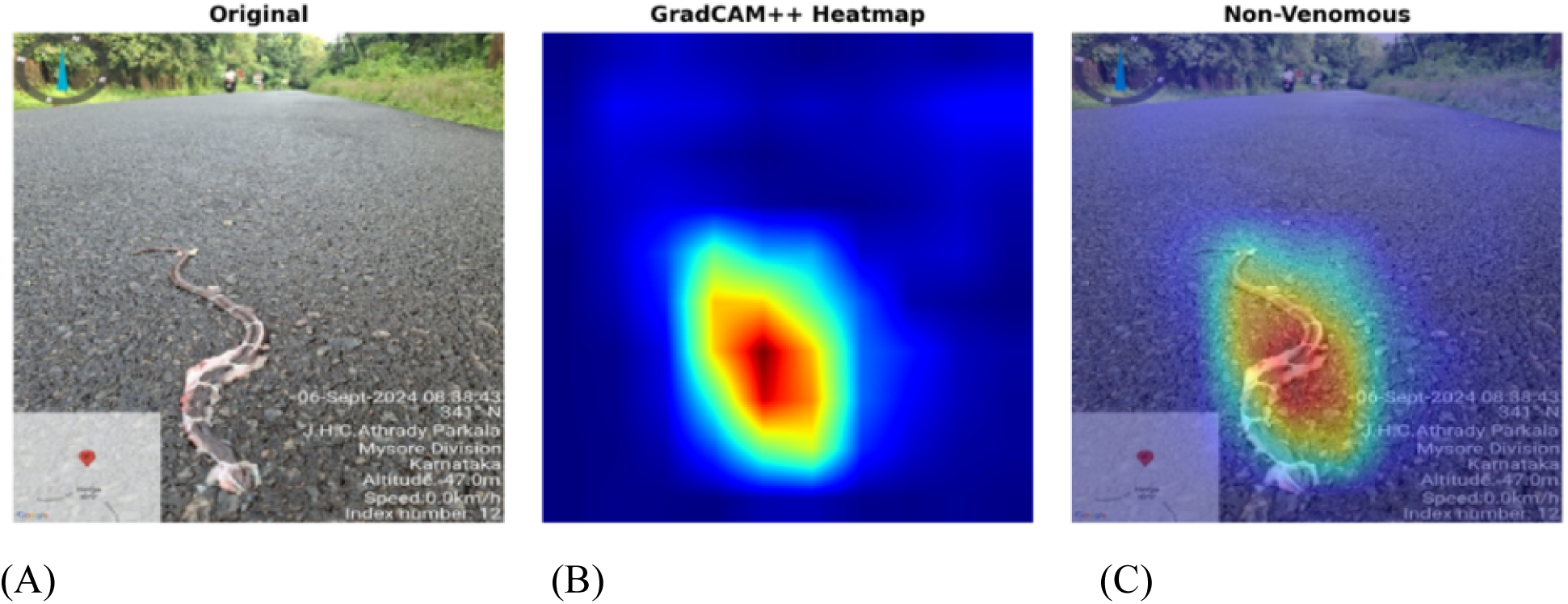
Non-Venomous Image: (A) Original image, (B) Gradcam image (C) Gradcam Overlay

As shown in Fig 8 (A) presents a venomous example. It shows the original image with the subject positioned in a complex natural environment. Fig 8 (B) depicts the GradCAM++ heatmap [16] where strong activations appear around key anatomical regions, particularly the head and upper body. Fig 8 (C) illustrates the overlay result. It demonstrates that the model focuses on biologically relevant features while reducing background influence.

**Fig 8:**
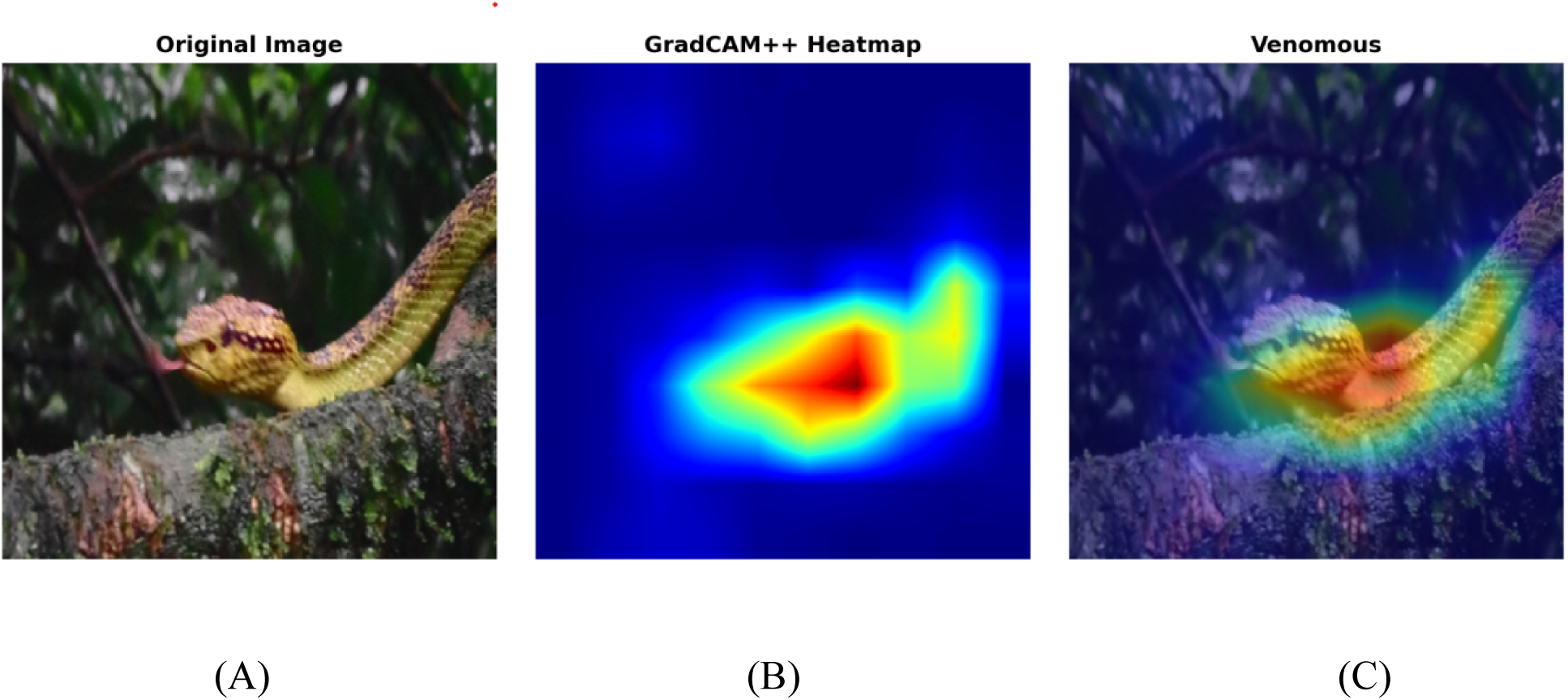
Venomous Image (A) original Image, (B) Gradcam Image (C) Gradcam Overlay

Overall, the visual explanations show that predictions rely on clear morphological features, supporting their understandability and reliability. Fig 9 (A) and 9 (B) show the web-based snake classification system. Users can upload an image, and the model predicts whether a snake is venomous or non-venomous, along with a confidence score. It shows the input image next to the result for clarity. A doctor or expert validation module allows for confirmation or changes to predictions, involving human oversight in sensitive clinical situations [19,20]

**Fig 9:**
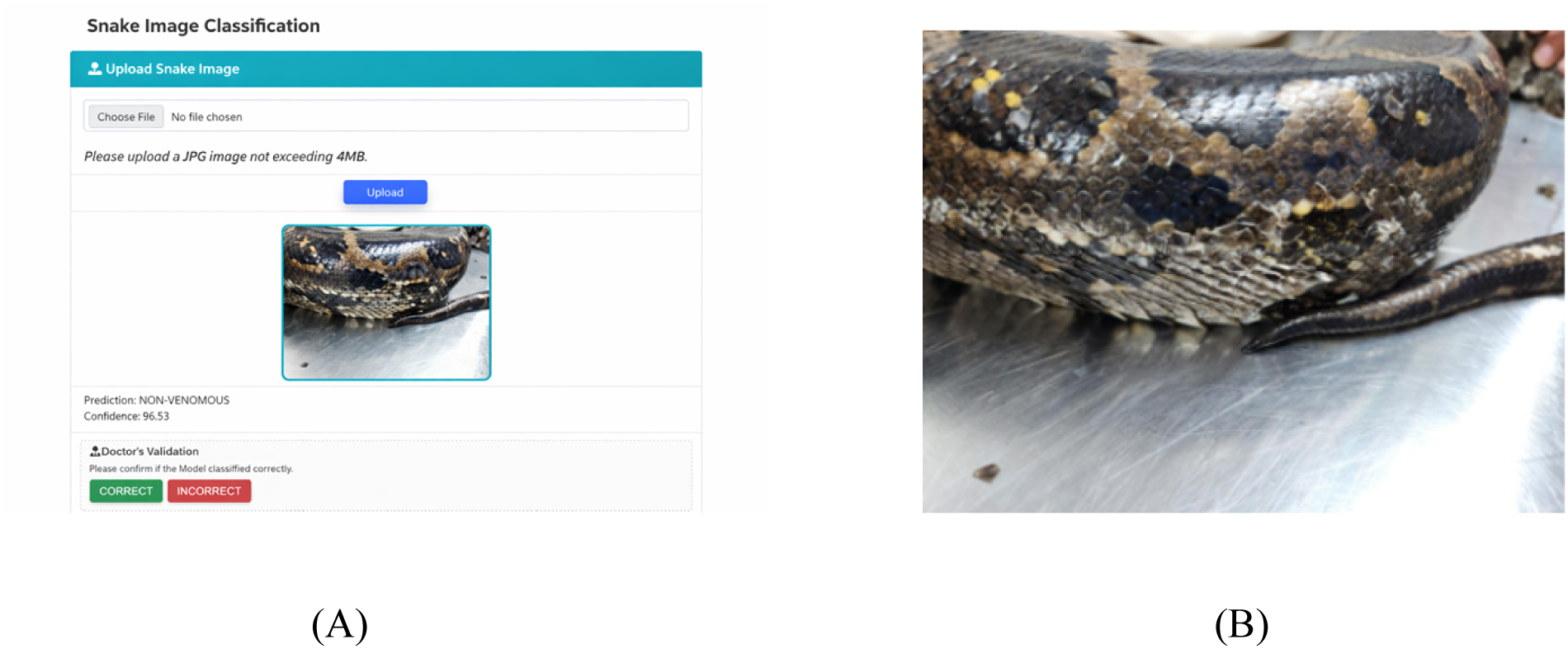
Deployment (A) Graphical User Interface (B) Snake Image being used to test

As shown in Figure 9 real-world image of the Indian Rock Python (Python molurus), which is non-venomous is classified correctly with 96.53% confidence, even though it looks similar to some venomous species.

Overall, the system offers a useful end-to-end solution that combines deep learning with expert validation. This is in line with new AI-assisted snake identification systems [11] [20]

## 4. Discussion

In real-world snakebite triage, delayed risk assessment can be life-threatening. The proposed ResNeXt-50 (32×4d) framework shows that it can be applied beyond benchmark evaluation to clinical decision-support situations.[15] It prioritizes venomous recall, uses meaningful feature learning, and validates predictions with Grad-CAM++ heatmaps and human reviews. This system balances performance with clinical responsibility. While it does not replace expert judgment, its high-confidence and reliable classification under realistic imaging conditions makes it a useful assistive tool for rural healthcare, field responders, and telemedicine workflows that need immediate risk assessment. As shown in Table 2, Rajabizadeh and Rezghi (2021) reported an accuracy of 93.16% using deep CNN transfer learning, with precision, recall, and F1-scores for multi-species classification.[21] However, their focus was on species-level recognition rather than binary clinical triage. Similarly, Iguernane et al. (2025) achieved an accuracy of 94% and an F1-score of 0.9586 in multi-class species identification, treating venom status as a species attribute rather than a separate medical risk classifier.[22] In contrast, this study targets binary venomous versus non-venomous classification for clinical triage support, achieving an accuracy of 97.64% and an F1-score of 0.9662. It also incorporates safety-oriented evaluation to reduce venomous false negatives, along with Grad-CAM++ interpretability and expert validation.

**Table 2:**
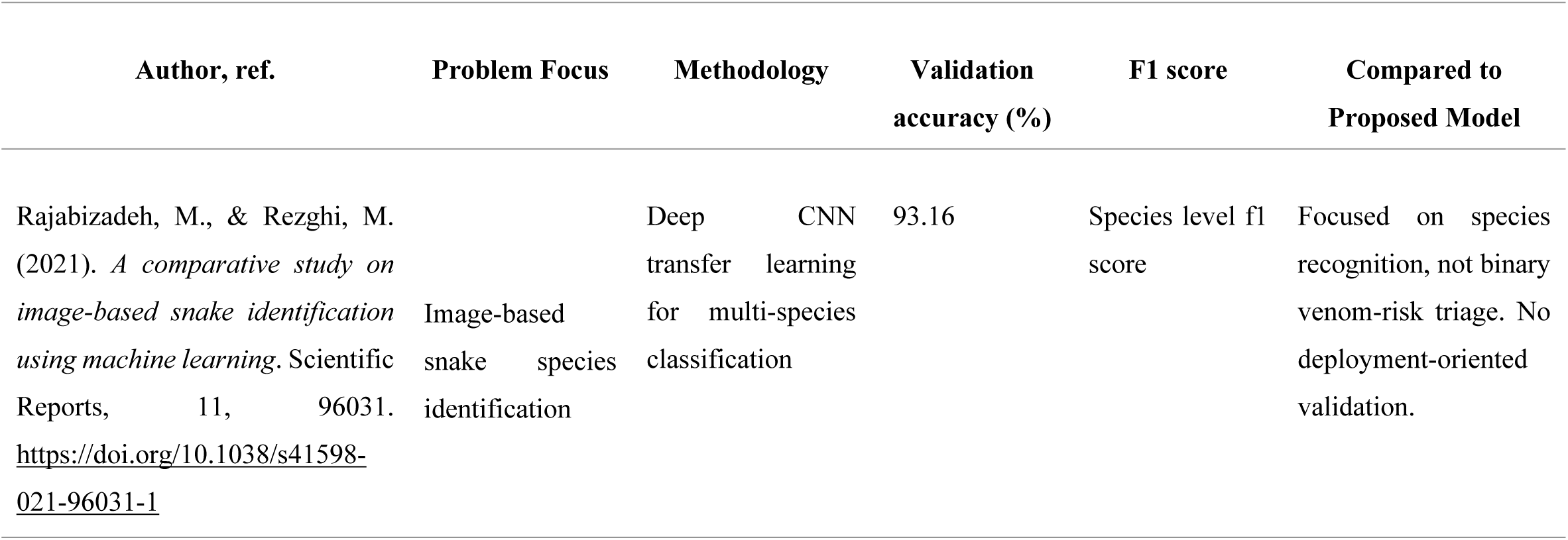
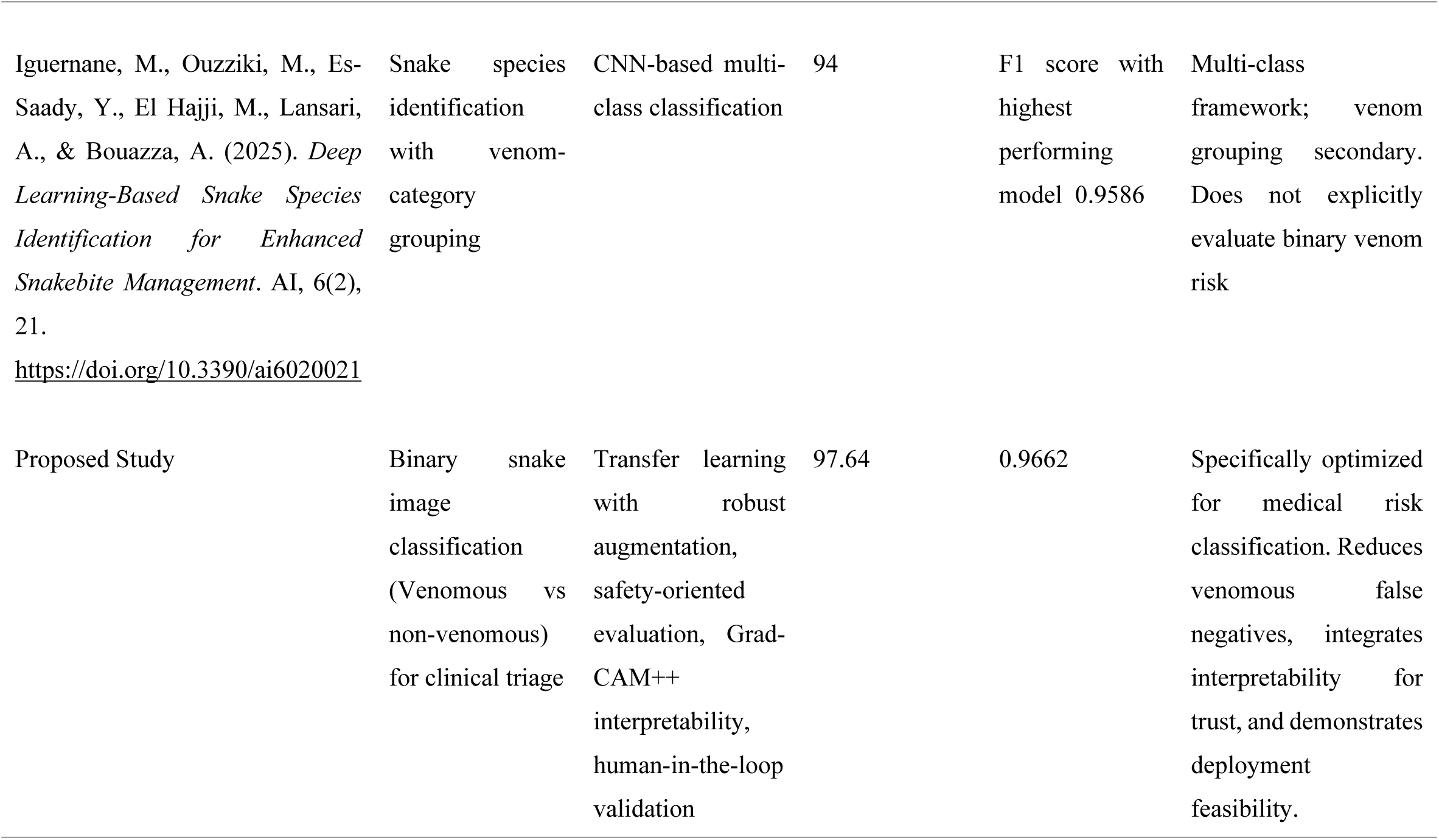
Comparative study of proposed work with the current literature.

### Role of AI Snake venomous snake Identifier in Snakebite registries

Snakebite registries are essential tools for epidemiological surveillance, outcomes tracking and policy developments in endemic regions in India. [1,23] Structured registry initiatives such as VENOMS registry in the Coastal Karnataka have demonstrated the feasibility of systematic documentation of envenomation cases across emergency departments. A persistent limitation in this system is the absence of standardized and objectively verified snake species classification. Species identification frequently depends on patient recall, informal local naming or non-standardized visual interpretation, poor quality images which may introduce reporting bias and misclassification.[24]. Misidentification of non-venomous species as venomous may lead to unnecessary administration of anti-snake venom, thereby exposing patients to avoidable risks like anaphylaxis, and under-recognition of venomous species may delay definitive treatment. Therefore, AI-assisted venomous snake identifier integrated into a registry framework can significantly enhance data fidelity.[24] By enabling real-time image upload and automated binary venom-risk classification and subsequent expert verification, registry entries can be supplemented with structured, reproducible risk labeling.

The VENOMS Helpline can serve as a triage and verification layer within this ecosystem. The VENOMS Helpline is a 24/7 telemedical support initiative by Kasturba Hospital (KH), Manipal, officially launched on 19th September 2025 as a collaborative program aimed at strengthening emergency response and clinical guidance for snakebite envenomation and other venomous exposures. The helpline is designed to support healthcare providers, emergency medical responders, and community members by facilitating rapid access to expert consultation during time-critical envenomation emergencies. When integrated with the VENOMS Helpline, the AI system will facilitate rapid preliminary venom-risk stratification, followed by structured expert review and verification to maintain clinical governance and decision-making integrity. This integrated model has immediate implications for rational ASV administration and real time triage decision-making, while also contributing to long term improvements in registry standardization, surveillance accuracy and continuous quality improvement within snakebite management systems.

### Role of AI Snake Identifier in primary and pre-hospital settings

Timely triage is critical in snakebite management, as delays in antivenom administration are associated with worse outcomes. [23] In rural and resource-limited regions, primary care centers and emergency medical services frequently serve as the first point of contact. However, expertise in snake identification is often unavailable at these levels of care. Integration into an EMS console platform allows paramedics to upload field-acquired images during patient transport. Real-time AI classification may assist in Early referral activation, pre-arrival antivenom preparation, resource mobilization at tertiary centers and Structured communication between peripheral facilities and referral hospitals. The VENOMS Helpline extends this functionality to telemedicine settings, enabling rural healthcare workers or community members to transmit images for rapid AI-assisted triage, followed by expert review. This integrated approach has immediate implications for improved prehospital decision-making and early system activation, while also contributing to long-term strengthening of referral pathways, documentation quality, and continuous quality improvement, serving as a bridge to healthcare access in settings with shortages of trained personnel and domain experts and may help reduce the overall economic burden associated with emergency snakebite care.

### Reliability and expert verification of images and syndromes associated with evidence provided

AI systems in healthcare require interpretability and oversight to ensure safe adoption. In this study, Grad-CAM++ visualization was incorporated to examine whether model predictions were based on anatomically meaningful features rather than spurious background artifacts. Activation maps demonstrated consistent focus on biologically relevant regions including Head morphology, Body contour, Scale patterns, and Relative head–body proportion. Such anatomical localization supports biological plausibility of learned features, which is a key requirement for reliable AI deployment in medicine. Image-based classification does not equate to envenomation severity. Clinical syndrome remains the gold standard determinant of management. Therefore, the developed web interface incorporates a human-in-the-loop validation module, allowing expert confirmation or override of predictions. This approach reflects established frameworks advocating supervised AI deployment in safety-critical contexts.

### Future directions: Cross regional and Global Generalizability

Snakebite epidemiology varies significantly across regions due to differences in distribution of species and venom composition.

Future directions include:

1. Multi center validation across diverse Indian states

1. Learning collaborations across institutions
2. Comparative evaluation against international snake classification datasets.
3. Prospective validation on patient-acquired smartphone images under real-world field conditions.

As a further step, integration of image-based classification with clinical syndrome profiling is recommended to enhance decision-support accuracy. Prior to this, additional model refinement with species-specific identification training will be undertaken to improve granularity beyond binary venom-risk stratification.

Such expansions align with global snakebite reduction strategies supported by the World Health Organization. Furthermore, region-specific performance benchmarking may inform policy decisions regarding AI-assisted triage systems globally.

### Ethical Consideration in AI assisted care

The deployment of artificial intelligence in clinical settings must align with core ethical principles to ensure safe, equitable, and trustworthy use. A foundational ethical framework rooted in the four principles of biomedical ethics - beneficence, non-maleficence, respect for autonomy, and justice is broadly applicable to medical AI systems and guides responsible implementation in healthcare practice [25]. Despite its potential, the clinical integration of AI raises important ethical concerns related to reliability, bias, transparency, and accountability.[25] Clear governance structures and defined responsibility attribution are essential to ensure that AI systems function within established clinical and ethical boundaries. Such safeguards are critical to maintaining patient safety and professional accountability in safety-critical medical contexts.

To mitigate these concerns, AI tools should be designed with interpretability and human oversight at their core, preserving clinician autonomy and accountability by ensuring that AI outputs remain advisory rather than prescriptive. Expert verification and human-in-the-loop validation help uphold non-maleficence by guarding against harmful decisions based on misclassification. Principles of justice require that AI systems be developed and tested with diverse data and governance structures that promote equitable access and prevent reinforcement of existing disparities, particularly in low-resource and underserved settings. By embedding these ethical considerations into the design and deployment of clinical AI, this study’s framework aims to uphold patient safety and trust while enhancing the utility and equity of AI-assisted snakebite management.

### 4.1 Study Limitations

1. The dataset, comprising 1,110 venomous and 384 non-venomous images prior to Augmentation, is relatively modest for a high-stakes clinical application. Additionally, all images were sourced from a single institution, limiting claims of geographic and demographic generalizability.
2. The current binary output does not extend to species-level identification or antivenom selection guidance, which would substantially increase clinical utility in practice [10]

Future work should focus on expanding the dataset to include underrepresented yet medically significant species, incorporating prospective real-world validation cohorts, developing multi-class species-level outputs to support antivenom selection, and exploring lightweight model distillation techniques for deployment on edge devices in connectivity-limited rural settings [23].

### 4.2 Conclusion

This study presents a clinically oriented, binary snake classification system trained on a curated Indian species dataset, offering a practical, interpretable, and robust AI-assisted solution to a critical unmet need in global snakebite management. While not a replacement for expert clinical judgment, the system’s anatomically grounded predictions under realistic imaging conditions suggest its potential as a scalable assistive tool for rural healthcare workers, emergency responders, and telemedicine platforms operating in snakebite-endemic regions. Further prospective validation and multi-center evaluation are necessary to establish generalizability and confirm performance prior to real-world clinical implementation.

## Data Availability

All data will be made available fully on acceptance, The Manuscript details all relevant information.

## Acknowledgements

We would like to thank Department of Emergency Medicine, Centre for Wilderness Medicine, KMC Manipal, Centre for Software Development and our colleagues for their support.

